# Omega-3 fatty acids and intracranial aneurysms: a Mendelian randomization study

**DOI:** 10.1101/2024.10.11.24315356

**Authors:** Dachao Wei, Xiheng Chen, Siming Gui, Jia Jiang, Yuan Gao, Jun Lin, Dingwei Deng, Wei You, Jian Lv, Yudi Tang, Ting Chen, Shu Yang, Hengwei Jin, Yuhua Jiang, Peng Liu, Hongfei Tai, Xinke Liu, Huijian Ge, Ming Lv, Fangang Meng, Youxiang Li

**Affiliations:** Beijing Neurosurgical Institute, Capital Medical University, Beijing, China; Department of Neurosurgery, Beijing Chaoyang Hospital, Capital Medical University, Beijing, China; Department of Interventional Radiology, Beijing Chaoyang Hospital, Capital Medical University, Beijing, China; chool of Biomedical Engineering, Capital Medical University, Beijing, China; Department of Neurosurgery, Beijing Tiantan Hospital, Beijing, China; Department of Neurology, Beijing Tiantan Hospital, Beijing, China; Chinese Institute for Brain Research, Beijing, China

**Author notes:** These authors contributed equally to this work (Dachao Wei, Xiheng Chen, Siming Gui). Corresponding author: Youxiang Li, MD, Beijing Neurosurgical Institute, Capital Medical University, No. 119 South Fourth Ring West Road, Fengtai District, Beijing 100050, China; Fangang Meng, MD, Beijing Neurosurgical Institute, Capital Medical University, No. 119 South Fourth Ring West Road, Fengtai District, Beijing 100050, China; Ming Lv, MD, Beijing Neurosurgical Institute, Capital Medical University, No. 119 South Fourth Ring West Road, Fengtai District, Beijing 100050, China.

**Keywords:** Intracranial aneurysm, Docosahexaenoic acid, Aneurysmal subarachnoid haemorrhage, Polyunsaturated fatty acid, Mendelian randomization

## Abstract

**Introduction:** Recent studies suggest that omega-3 polyunsaturated fatty acids (PUFAs) supplementation benefits for cardiovascular disease and abdominal aortic aneurysms, but its role in intracranial aneurysms (IAs) remains unclear. This study evaluates the effect of omega-3 PUFAs on IAs.

**Patients and methods:** A two-sample Mendelian Randomization study (MR) was conducted to examine the associations between omega-3 PUFAs and IAs, as well as aneurysmal subarachnoid hemorrhage (aSAH). The largest genome-wide association study dataset was used for primary analysis, with replication using independent sources. Two-step MR was used to evaluate the mediating effects of nine aneurysm risk factors and 91 inflammatory cytokines.

**Results:** Higher genetically predicted levels of total omega-3, omega-3 percentage, and docosahexaenoic acid (DHA) were associated with reduced risks of IAs (combined OR 0.88, 95% CI 0.83-0.94, P<0.001; 0.86, 0.81-0.92, P<0.001; 0.84, 0.78-0.90, P<0.001, respectively) and aSAH (0.85, 0.79-0.91, P=0.009; 0.84, 0.75-0.93, P<0.001; 0.80, 0.71-0.90, P<0.001, respectively). Genetically predicted eicosapentaenoic acid level showed a tendency to increase risk (1.05, 1.01-1.10, P=0.022 for IAs; 1.11, 0.96-1.28, P=0.160 for aSAH). Mediation analysis revealed diastolic blood pressure (DBP) and tumor necrosis factor-related apoptosis-inducing ligand (TRAIL) mediated DHA’s effects on IAs (proportion mediated 8.5%, P=0.019; 25.2%, P=0.049) and aSAH (5.7%, P=0.024; 27.5%, P=0.041).

**Conclusions:** Our study suggests that omega-3 PUFAs, particularly DHA, are associated with a reduced risk of IAs and aSAH. This association may be partially mediated by DBP and TRAIL.

## Introduction

Intracranial aneurysms (IAs) affect approximately 3% of the global population^1^. When ruptured, they often cause aneurysmal subarachnoid hemorrhage (aSAH), with high morbidity and mortality rates^2^.

Despite the low rupture risk of unruptured intracranial aneurysms (UIAs) (∼0.25%)^3^, their unpredictable nature poses a clinical challenge^3, 4^. Physicians must balance the benefits and risks of preventive surgery, as rupture prediction tools are inadequate^4^. Therefore, interest is growing in non-invasive pharmacological interventions^5^.

Inflammation is a key driver of IA pathogenesis, contributing to arterial wall weakening and rupture risk^6, 7^. Targeting inflammatory pathways could be a viable therapeutic approach. Omega-3 fatty acids, particularly eicosapentaenoic acid (EPA) and docosahexaenoic acid (DHA), are well-known for their anti-inflammatory properties^8, 9^ and have shown promise in reducing inflammation and oxidative stress in other vascular diseases, such as abdominal aortic aneurysms (AAAs)^10, 11^. This raises the possibility that these fatty acids could play a protective role in the context of IAs.

However, observational studies are often confounded, limiting causal inference. Mendelian randomization (MR) offers a robust alternative by using genetic variants to assess causality^12^. This study aims to use MR to explore the effects of omega-3 fatty acids on IA development and rupture, providing insight into their potential as preventive interventions.

### Patients and methods

We conducted two-sample MR to assess the role of omega-3 fatty acids in IA formation and rupture, using summary-level data from recent Genome-Wide Association Studies (GWASs). Mediation analysis explored underlying mechanisms. The study follows STROBE-MR guidelines, with ethics approval obtained in all original studies.

### Data sources

Data sources are summarized in Table S1. We selected GWAS datasets for exposures, mediators, and outcomes based on strict criteria to avoid overlap and maximize sample size.

We analyzed four omega-3 indices: total omega-3 concentration, omega-3 percentage (omega-3%), plasma DHA, and plasma EPA. Data for the first three indices were from the UK Biobank, with omega-3 levels measured via NMR spectroscopy in 115,078 participants^13^. EPA data were obtained from the CHARGE Consortium, including 8,866 European adults^14^.

Outcome data were sourced from the largest GWAS meta-analysis by Bakker et al., which combined 23 cohorts where IAs were diagnosed via angiography^15^. The primary analysis used stage 1 and European ancestry aSAH data from Bakker et al., while replication analyses used data from FinnGen R9 and GWAS datasets, including Sakaue *et al*.^16^ and other datasets from Bakker *et al*. Detailed information regarding exposure and outcome data is provided in the **Supplementary Methods**.

### Genetic instruments extraction

IVs strongly associated with exposures, (P<5×10^−8^ for total omega-3, omega-3%, DHA, P<1×10^−5^ for EPA) were selected from the GWAS data and clumped to remove variants in linkage disequilibrium (r^2^<0.001 and clump distance>10,000 kb). We calculated R^2^ and F-statistics for the selected IVs as previously described^17^. Weakly associated SNPs (F-statistic<10) were discarded. SNPs satisfying the hypotheses were obtained from the outcome datasets, and if unavailable, proxy SNPs were identified at a linkage disequilibrium R^2^>0.8. Exposure and outcome data were harmonized to ensure consistent allele alignment.

### Two-sample MR analysis

In MR, adherence to three key assumptions is imperative for valid results (**Figure 1**). The primary MR analysis in this study was the inverse variance-weighted (IVW) method. Additional analyses were conducted using MR-Egger, weighted median, and weighted mode methods. These methods offer robustness in the presence of invalid instruments but with reduced statistical power^18^. The statistical methods used in this study are detailed in **Supplementary Methods**.

**Figure 1.**
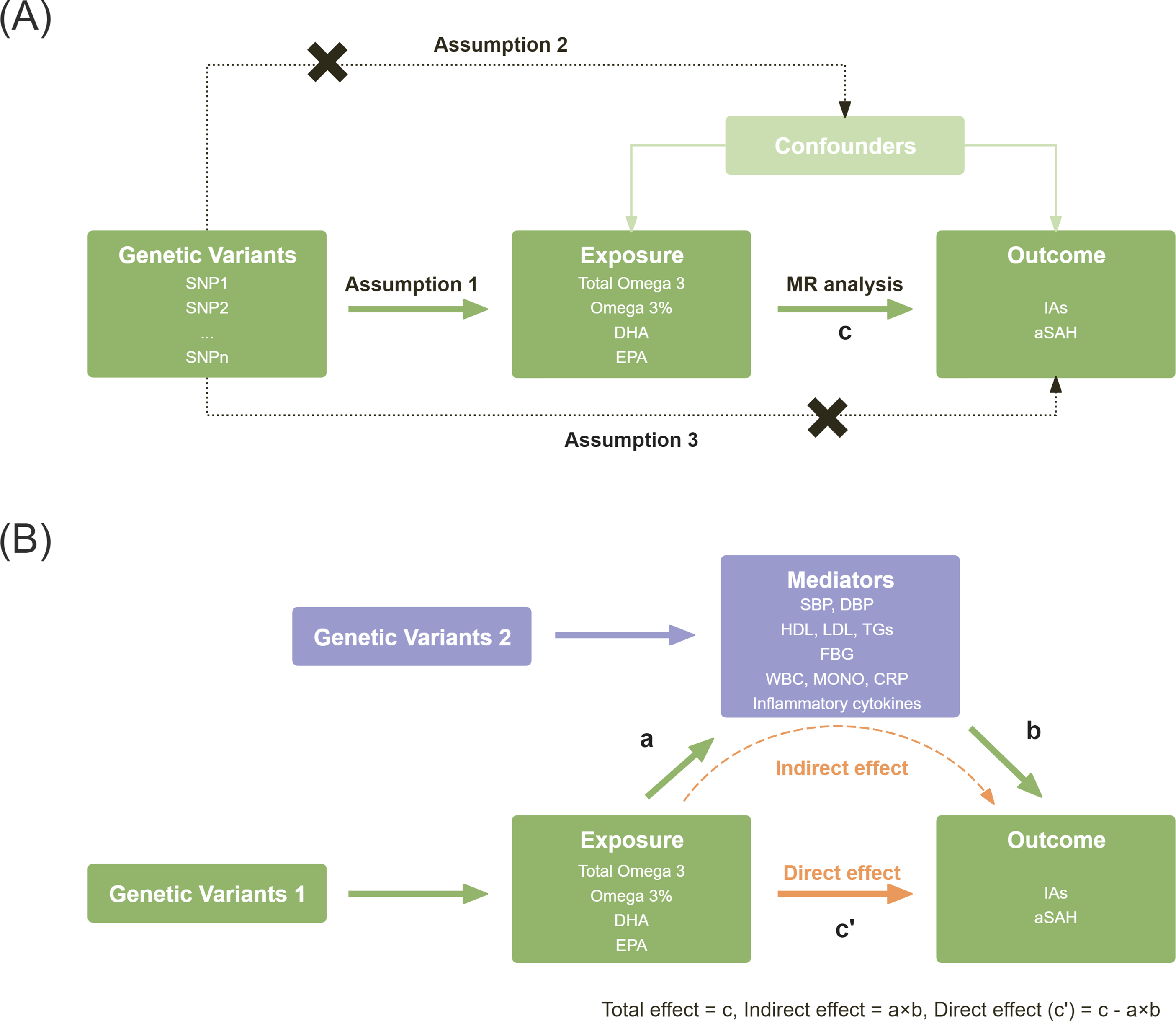
A: Diagram showing the key assumptions of two-sample Mendelian randomization. The total effect (c) is the overall effect of exposure on the outcomes. Mendelian randomization studies adhere to three fundamental hypotheses: 1) The instrumental variables (IVs) are associated with the exposure (Assumption 1). 2) IVs are independent of the confounding factors that confound the association of exposure and the outcome (Assumption 2). 3) IVs influence the outcome only through the exposure (Assumption 3). B: Diagram showing Mendelian randomization mediation analysis. The indirect effect (a × b) is the effect of exposure on the outcome through the mediator. The direct effect (c’) is the effect of exposure on the outcome without being mediated by any intermediate variable (c − a × b). The mediated proportion is calculated as c’ divided by c. Abbreviations: SNP, single nucleotide polymorphism; Abbreviations: DHA, docosahexaenoic acid; EPA, eicosapentaenoic acid; IAs, intracranial aneurysms; aSAH, aneurysmal subarachnoid hemorrhage; LDL-C, low-density lipoprotein cholesterol; HDL-C, high-density lipoprotein cholesterol; TGs, triglycerides; SBP, systolic blood pressure; DBP, diastolic blood pressure; FBG, fasting blood glucose; WBC, white cell count; MONO monocyte count; CRP, C-reactive protein.

### Replication and Sensitivity analyses

Replication MR analyses were conducted using data from Sakaue et al., FinnGen, and Bakker et al. We combined primary and replication results using random-effects meta-analysis for increased precision. Sensitivity analyses included MR-Egger intercept tests to detect horizontal pleiotropy and Cochran’s Q for heterogeneity. A leave-one-out analysis was conducted to check if any single SNP was driving the causal signal.

### Mediation analysis

Mediation analyses were conducted to assess mechanisms underlying the effect of exposure on outcomes. Based on a comprehensive literature review, we identified potential mediators including blood pressure (systolic blood pressure [SBP], diastolic blood pressure [DBP]), blood lipids (low-density lipoprotein [LDL], high-density lipoprotein [HDL], triglycerides [TGs], blood glucose (fasting blood glucose [FBG]), inflammation markers (white blood cell count [WBC], monocyte count [MONO], C-reactive protein [CRP]), and inflammatory cytokines.

We collected the largest genome-wide association study dataset to date for these mediators, ensuring European ancestry and no population overlap with exposures and outcomes (**Table S1**). Genetic variants were extracted according to the previously mentioned method.

A two-step MR approach was used to evaluate the presence of a mediating effect. This involved using two-sample MR to calculate the total effect from exposure to outcomes, the effect of exposure on mediators, and the effect of mediators on outcomes, followed by the calculation of the mediating effect. The Sobel test assessed mediation significance, with 95% confidence intervals (CIs) determined using RMediation^19^.

### Statistical analysis

All analyses were performed using R (version 4.2.1) and the TwoSampleMR package (version 0.5.6). Bonferroni correction (P<0.025[0.05/2]) was applied for multiple testing.

## Results

### Genetic variants

We included 52, 41, 48, and 50 SNPs strongly associated with total omega-3, omega-3%, DHA, and EPA, respectively. IVs had F-values from 19.4 to 4825.5, indicating minimal weak instrument bias **(Table S2)**.

### Effect of Omega-3 PUFAs on IAs

As shown in **Figure 2**, the primary MR analysis demonstrated that genetically predicted total omega-3, omega-3%, and DHA were associated with a reduced risk of IAs (odds ratio [OR_IVW_] 0.84, 95%CI 0.74–0.96, P=0.011; OR_IVW_ 0.81, 95%CI 0.70–0.94, P=0.004; OR_IVW_ 0.78, 95%CI 0.68–0.90, P<0.001, respectively). However, genetically predicted EPA level tended to associate with increased risk of IAs (OR_IVW_ 1.16, 95%CI 0.98–1.39, P=0.092).

**Figure 2.**
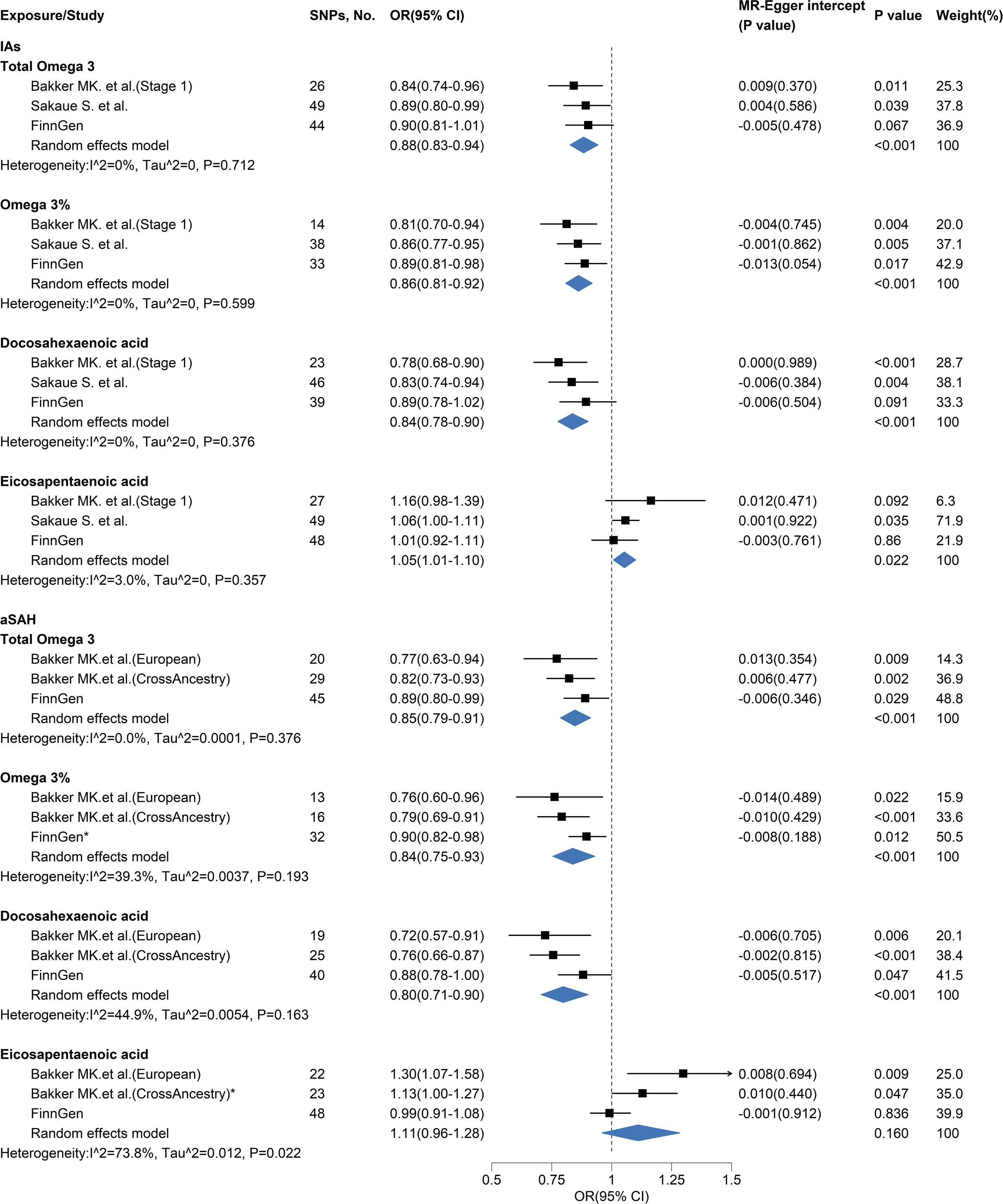
Univariable effects of omega-3 fatty acids on intracranial aneurysms and aneurysmal subarachnoid hemorrhage. Abbreviations: OR, odds ratio; CI, confidence interval; IAs, intracranial aneurysms; aSAH, aneurysmal subarachnoid hemorrhage.

In the replication and meta-analyses, the results were largely consistent with the main analysis, with EPA showing an increased risk (ORmeta 1.05, 95%CI 1.01–1.10, P=0.022) (**Figure 2**).

### Effect of Omega-3 PUFAs on aSAH

For aSAH, the primary MR analysis of omega-3 PUFAs yielded similar results: total omega-3, omega-3%, and DHA were associated with a reduced risk of aSAH (OR_IVW_ 0.77, 95%CI 0.63–0.94, P=0.009; OR_IVW_ 0.76, 95%CI 0.60–0.96, P=0.022; OR_IVW_ 0.72, 95%CI 0.57–0.91, P=0.006), whereas EPA was associated with an increased risk of aSAH (OR_IVW_ 1.30, 95%CI 1.07–1.58, P=0.009) (**Figure 2**).

These associations remained consistent and significant in both the replication MR analysis and meta-analysis. However, the EPA analysis with the FinnGen dataset showed a different direction of effect but was not statistically significant (**Figure 2**).

### Sensitivity analyses

Results for all omega-3 PUFAs analyses were consistent across different MR methods (**Table S3**). Heterogeneity was substantial in most analyses, but there was no evidence of directional pleiotropy (**Figure 2** and **Tables S4 and S5**). The leave-one-out analysis also did not suggest that a single SNP could alter the direction of the effect, except for some SNPs in EPA (**Tables S6 and S7**).

### Mediation analysis for established aneurysm risk factors

In our MR analysis, DHA had a lower OR for both IA and aSAH compared to total omega-3 and omega-3%, while EPA had an OR greater than 1. These results suggest that different omega-3 fatty acids may have varying effects, with DHA potentially playing a major role in reducing the risk of these conditions. To explore the potential mechanisms underlying the protective effects of DHA, we conducted a mediation analysis involving aneurysm risk factors.

In the analysis of the mediating effect of DHA, genetically determined DHA concentrations were inversely associated with DBP (OR_IVW_ 0.98, 95%CI 0.97–1.00, P=0.012) and MONO (OR_IVW_ 0.99, 95%CI 0.97–1.00, P=0.024). Regarding the impact of these mediators on outcomes, SBP, DBP were correlated with an increased risk of IAs (OR_meta_ 2.73, 95%CI 2.14–3.48, P<0.001; OR_meta_ 2.36, 95%CI 1.82–3.06, P<0.001), as well as an increased risk of aSAH (OR_meta_ 2.88, 95%CI 2.44–3.40, P<0.001; OR_meta_ 2.50, 95%CI 1.89–3.31, P<0.001). MONO is associated with an increased risk of aSAH (OR_meta_ 1.12, 95%CI 1.03–1.21, P=0.006), while there was a tendency to increase the risk of IAs (OR_meta_ 1.08, 95%CI 1.00–1.16, P=0.052). However, FBG and CRP were inversely associated with IAs and aSAH (**Figure 3**).

**Figure 3.**
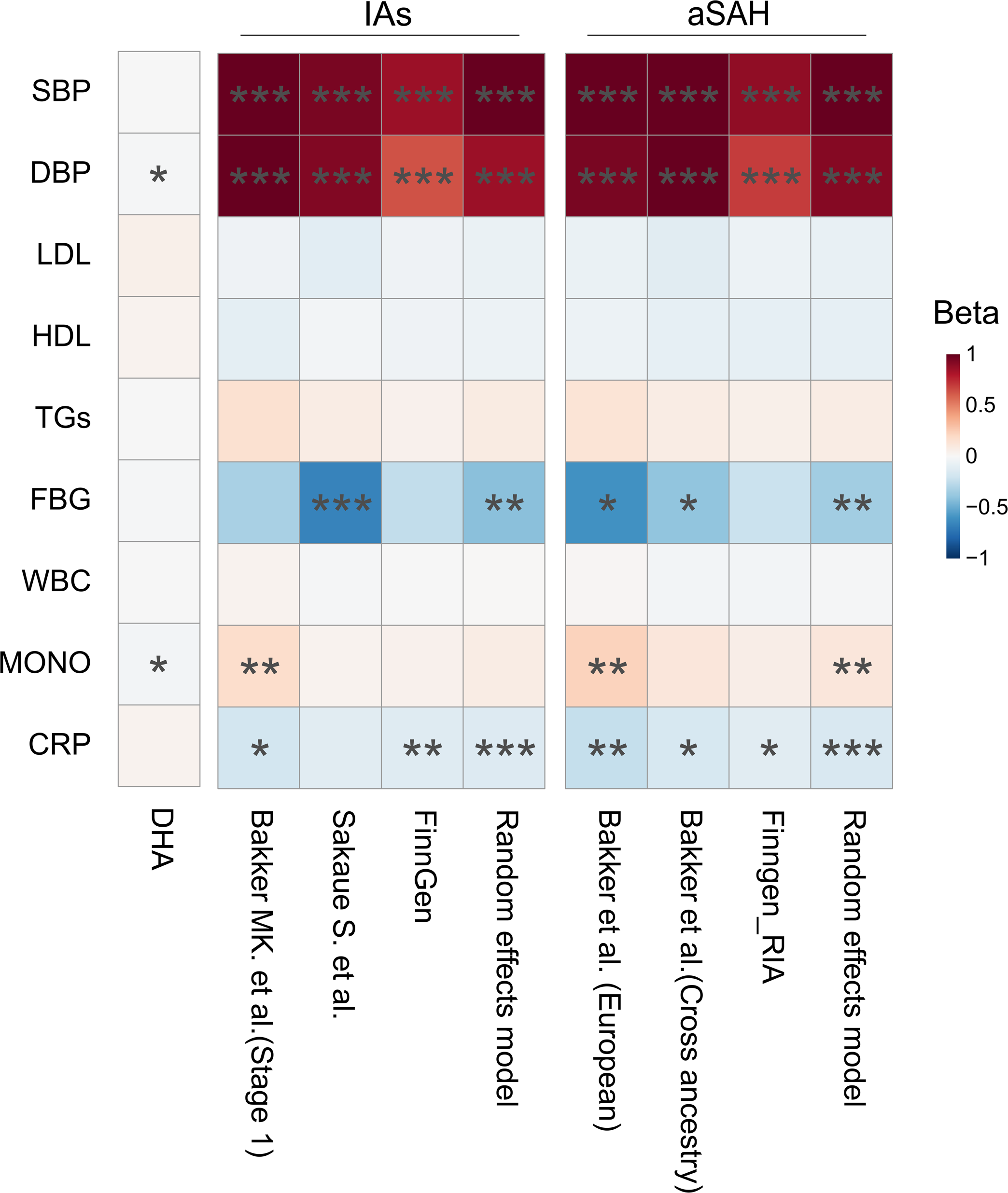
Mediation analysis of docosahexaenoic acid, aneurysm risk factors, and diseases. The results are depicted as beta coefficients, representing variations in different exposures (y-axis) across multiple data sources (x-axis). The associations are color-coded: blue boxes indicate decreases, and red boxes indicate increases in the risk of outcomes. Statistical significance is marked with asterisks: *** for P<0.001, ** for P<0.01, and * for P<0.05. Abbreviations: IAs, intracranial aneurysms; aSAH, aneurysmal subarachnoid hemorrhage; DHA, docosahexaenoic acid; LDL-C, low-density lipoprotein cholesterol; HDL-C, high-density lipoprotein cholesterol; TGs, triglycerides; SBP, systolic blood pressure; DBP, diastolic blood pressure; FBG, fasting blood glucose; WBC, white cell count; MONO monocyte count; CRP, C-reactive protein.

Using two-step MR, we observed an indirect effect of DHA concentrations on IAs and aSAH through DBP (OR 0.98, 95%CI 0.96–1.00, P=0.019; OR 0.98, 95%CI 0.97–1.00, P=0.024), with a mediated proportion of 8.4% and 5.7%, respectively. (**Figure 3** and **Table S30**).

In the sensitivity analysis, these effects in replication and meta-analysis are consistent, with no evidence of horizontal pleiotropy. However, most analyses exhibit significant heterogeneity (**Figure 3** and **Tables S8-S21**).

### Mediation analysis for inflammatory cytokines

Our research suggests that DBP plays a part in the intermediary effect of DHA on IAs and aSAH, but the effect is limited (8.4% and 5.7%). Given that inflammatory cytokines play a key role in IA formation and rupture, we further examined whether 91 different inflammatory cytokines mediate the protective effects of DHA.

Genetically determined DHA concentrations were significantly negatively associated with 24 inflammatory cytokines, with the most significant being tumor necrosis factor-related apoptosis-inducing ligand (TRAIL) (OR_IVW_ 0.79, 95%CI 0.72–0.87, P<0.001), (**Figure 5**). In the primary analysis of IAs, five cytokines were associated with increased risk, with TRAIL (OR_IVW_ 1.30, 95%CI 1.06–1.59, P=0.012) being the most significant (**Figure 5**). Similarly, in the primary analysis of aSAH, TRAIL (ORIVW 1.35, 95%CI 1.05–1.73, P=0.017) was the most significant cytokine (**Figure 5**).

Two-step MR revealed an indirect effect of DHA on IAs and aSAH through TRAIL (OR 0.94, 95%CI 0.88–1.00, P=0.049; OR 0.91, 95%CI 0.84–1.00, P=0.041), with mediated proportions of 25.2% and 27.5%, respectively. No evidence of heterogeneity or pleiotropy was found (**Figure 4, Tables S22-S29)**. Although the direction of effect was consistent in replication and meta-analysis, it was not statistically significant (**Figure 5**).

**Figure 4.**
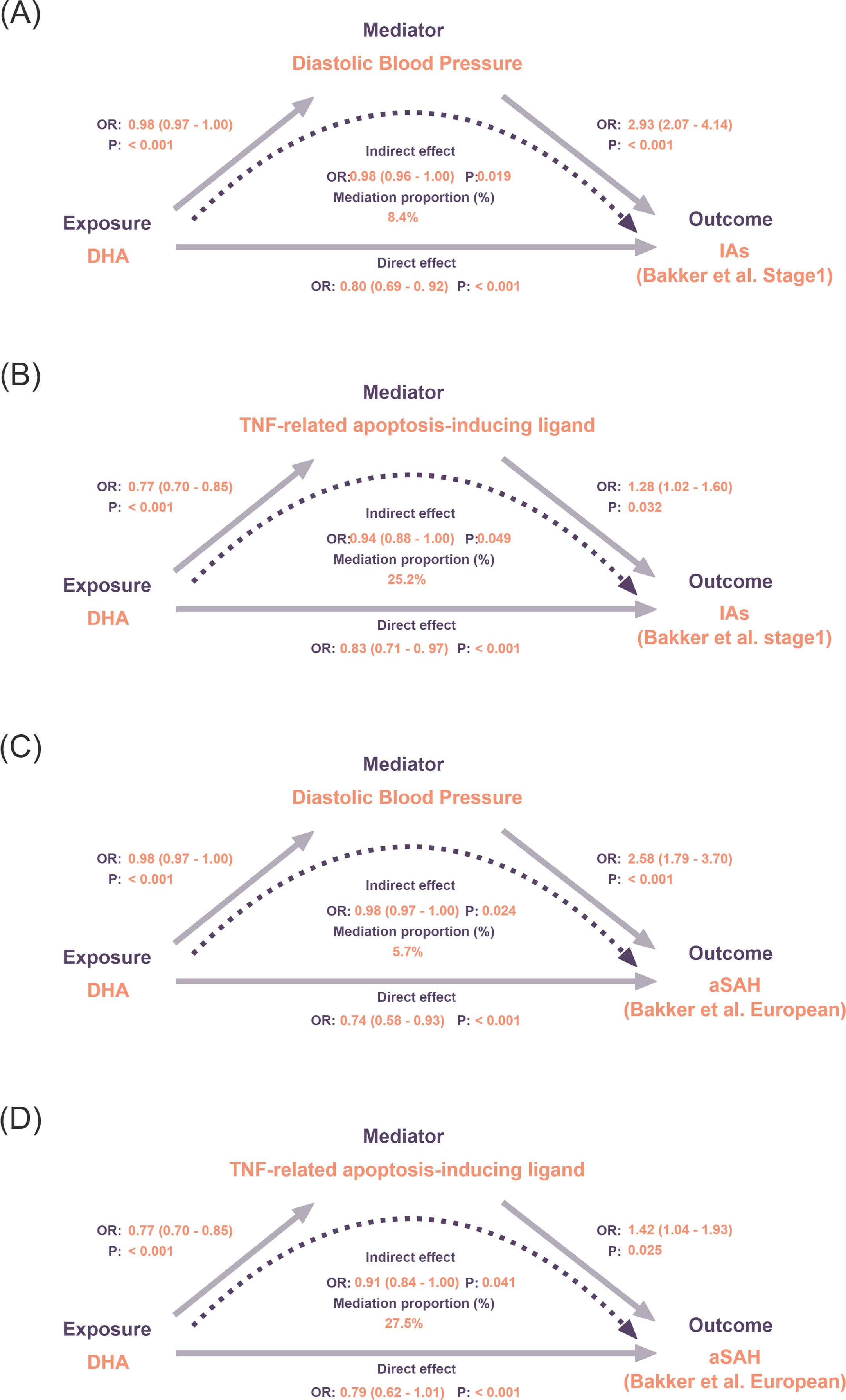
Summary of mediating pathways of docosahexaenoic acid (DHA)–mediators–diseases identified in this study. (A) The diastolic blood pressure mediated the effect of DHA on intracranial aneuysms. (B) tumor necrosis factor-related apoptosis-inducing ligand mediated the effect of DHA on intracranial aneuysms. (C) The diastolic blood pressure mediated the effect of DHA on aneurysmal subarachnoid hemorrhage. (D) tumor necrosis factor-related apoptosis-inducing ligand mediated the effect of DHA on aneurysmal subarachnoid hemorrhage. Abbreviations: IAs, intracranial aneurysms; aSAH, aneurysmal subarachnoid hemorrhage; DHA, docosahexaenoic acid; TNF, tumor necrosis factor.

**Figure 5.**
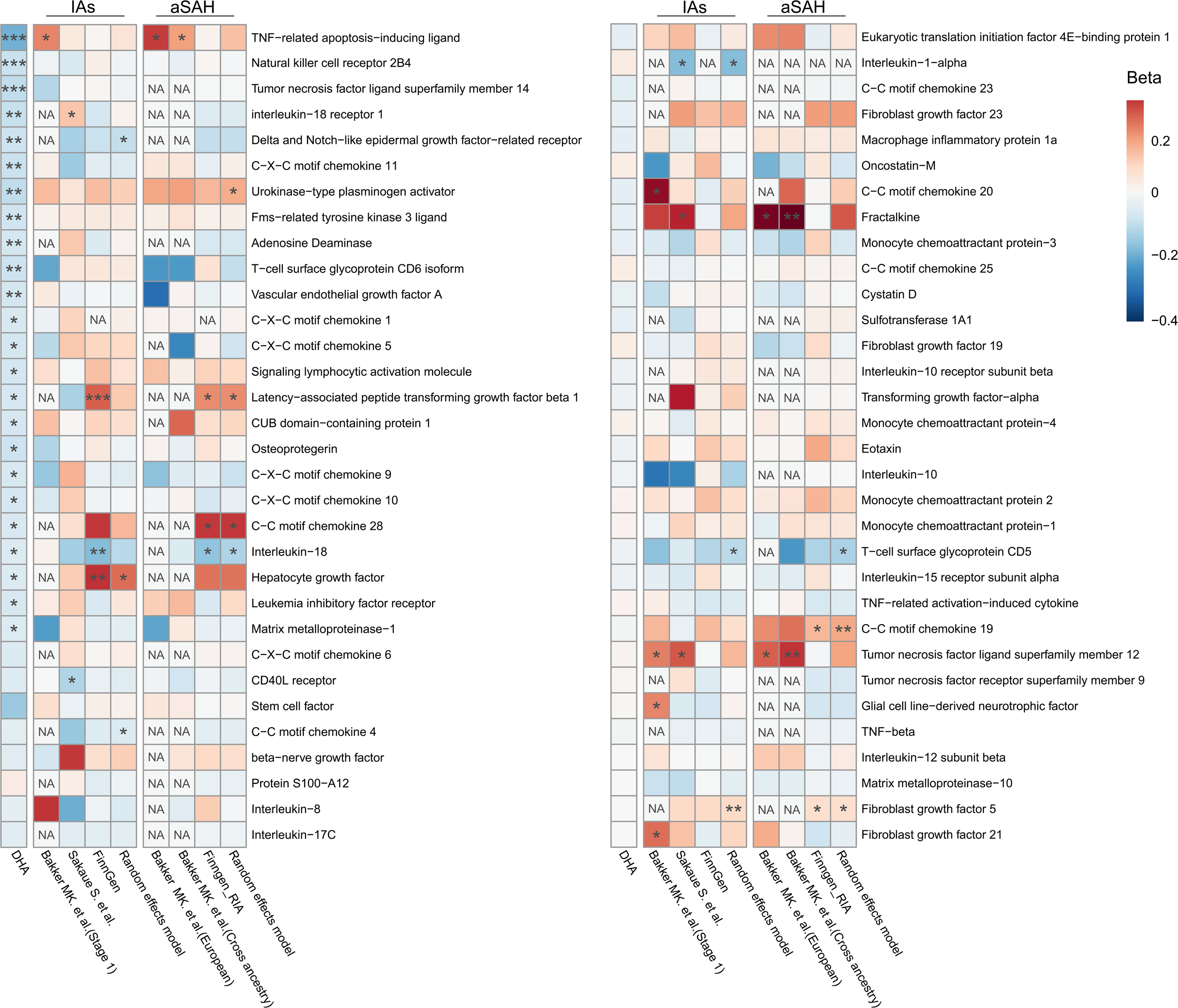
Mediation analysis of docosahexaenoic acid, 91 inflammatory cytokines, and diseases. The first column shows the influence of docosahexaenoic acid on inflammatory cytokines, while the second and third columns show the influence of inflammatory cytokines on intracranial aneurysms and aneurysmal subarachnoid hemorrhage, respectively. Results are presented as beta coefficients representing the variation in different cytokines (y-axis) across multiple data sources (x-axis). The associations are color-coded: blue boxes indicate decreases, red boxes indicate increases, and white boxes indicate no change in the risk of outcomes. Statistical significance is marked with asterisks: *** for P<0.001, ** for P<0.01, and * for P<0.05. NA indicates results not available due to a lack of genetic variants for conducting analysis. Abbreviations: IAs, intracranial aneurysms; aSAH, aneurysmal subarachnoid hemorrhage; DHA, docosahexaenoic acid.

Replication and meta-analysis found several other potential intermediary effects (urokinase-type plasminogen activator, latency-associated peptide transforming growth factor beta 1, C-C motif chemokine 28, interleukin-18, and hepatocyte growth factor) (**Figure 5**). However, all were marginal in the Sobel test (**Table S30**). These results underscore the need for larger studies to determine the influence of these inflammatory cytokines on IAs and aSAH.

## Discussion

### Principal findings

This study found that omega-3 PUFAs, especially DHA concentrations are associated with reduced risk of IAs (OR_IVW_ 0.78, 95%CI 0.68–0.90, P<0.001) and aSAH (OR_IVW_ 0.72, 95%CI 0.57–0.91, P=0.006). In contrast, increased concentrations of EPA were associated with a higher risk of both IAs and aSAH, suggesting that DHA may be the primary protective component among omega-3 fatty acids. Mediation analysis indicated that 8.4% and 5.7% of DHA’s protective effect was mediated by DBP. However, the role of inflammatory cytokines is inconsistent across different datasets. TRAIL may partially mediate the effect of DHA on IAs and aSAH.

### Comparison with other studies

The effects of omega-3 PUFAs on IAs and aSAH have not been well established in the literature, with limited studies available on this topic. Abekura et al.^8^ found that EPA significantly suppressed IA size and macrophage infiltration in rats. However, our Mendelian randomization analysis shows that EPA increased the risk of IAs and aSAH. This discrepancy may stem from differences between the long-term physiological levels of EPA reflected in MR and the effects of short-term, ex vivo EPA supplementation.

Several studies have examined the effects of omega-3 PUFAs on AAA development, with mixed results^11, 20–24^. In vivo studies, such as those by Yoshihara et al.^22^ and Kavazos et al.^23^, showed that DHA supplementation reduced AAA formation and inflammatory markers. Conversely, Aikawa et al.^24^ found an association between lower EPA levels and larger AAAs in patients, while DHA levels showed no correlation. This inconsistency may arise from differences in the form, dosage, and duration of omega-3 supplementation, underscoring the complex mechanisms of different PUFAs on aneurysm diseases.

### Possible mechanisms

In the mediation analysis, our study suggests that 8.4% and 5.7% of DHA’s protective effect against IAs and aSAH could be mediated by its impact on DBP. This finding adds a new dimension to the understanding of DHA’s role in vascular health, particularly its ability to lower blood pressure. While the blood pressure-lowering effects of omega-3 PUFAs have been explored extensively in prior research^25^, our study is among the first to specifically link DHA’s influence on blood pressure to a reduced risk of IAs and aSAH. The regulation of blood pressure by DHA occurs through both endothelium-dependent mechanisms, such as increasing nitric oxide availability, and endothelium-independent pathways, including inhibition of calcium inflow in arterial smooth muscle cells^26, 27^. This blood pressure modulation may explain the reduced risk of IAs and aSAH in patients with elevated DHA levels, particularly given the well-established relationship between hypertension and IAs^28^.

Previous studies have suggested that the anti-inflammatory effects of omega-3 PUFAs may help prevent the development of IAs. Our study supports these findings. We observed that genetically predicted DHA levels were associated with lower levels of monocytes and 24 inflammatory cytokines. Notably, TRAIL may mediate the protective effect of DHA on IAs and aSAH.

TRAIL is a transmembrane protein expressed in vascular smooth muscle, endothelial, and immune cells, known to induce apoptosis by binding to death receptors^29, 30^. Its role in IA progression may be similar to what is observed in AAAs, where it promotes apoptosis in smooth muscle cells and increases extracellular matrix degradation^31^.

However, the relationship between DHA and TRAIL remains understudied We speculate that DHA may reduce oxidative stress, thereby decreasing apoptotic inducers such as TRAIL. Under physiological conditions, arterial wall cells are frequently exposed to pathological stimuli, producing oxygen free radicals such as O2•− and H2O2, which increase oxidative stress. The expression of pro-apoptotic proteins like TRAIL may increase under the excessive oxidative stress, triggering apoptosis through intrinsic or extrinsic pathways^32, 33^. This process contributes to aneurysm formation or rupture^33^. DHA may counteract oxidative stress by activating the Keap1-Nrf2 pathway, promoting the production of endogenous antioxidants like glutathione peroxidases and superoxide dismutase^34, 35^. This enhances cellular defenses and reduces apoptosis and apoptotic inducers such as TRAIL^36^. Future research should explore DHA’s role in modulating TRAIL activity in human and animal models of IAs.

In conclusion, our study suggests that omega-3 PUFAs, particularly DHA, may have a potential preventive effect on IAs and aSAH through various mechanisms. This compound could serve as an adjunct therapy to prevent the formation and rupture of IAs. It may offer a pleiotropic advantage over blood pressure reduction alone, especially for IA patients with normal blood pressure—a condition observed in a significant proportion of aneurysm patients (62.8% [931/2505] in the ICAN population^37^ and 59.4% [2410/4060] in ISUIA cohort^38^), where blood pressure may not significantly decrease. Future research should focus on clinical applications in human populations and explore the mechanisms of action in animal models.

### Limitations

Our study has some limitations. First, MR studies depend on the assumptions of relevance, independence, and exclusion-restriction. Although we minimized bias using strong genetic instruments and conducted sensitivity analyses, the assumptions cannot be fully confirmed.

Second, some outcomes included a small East Asian population, raising the potential for stratification bias. However, the likelihood of these genetic variants inducing bias in the same direction is low^39, 40^. Despite mixed-race samples, our findings may not be generalizable to other populations.

Third, genetic variants reflect lifelong exposure impacts, which cannot be directly applied to short-term drug effects but offer insights into long-term exposure, often challenging to assess in randomized controlled trials.

Finally, MR-derived effect sizes may not directly translate to therapeutic benefits due to differences in drug dosages and patient responses, requiring validation through clinical trials.

## Conclusion

This study, using Mendelian randomization, suggests a possible causal relationship between DHA and a reduced risk of IA development and rupture. These findings offer a potential new perspective on the clinical application of omega-3 fatty acids, particularly DHA, as a preventive intervention for high-risk patients who may not be suitable for surgery. Future clinical studies are needed to validate these results and further explore the role of omega-3 fatty acids in different populations.

## Data Availability

Summary statistics for omega-3 measurements are available through the UK Biobank (application required at https://www.ukbiobank.ac.uk/) and the CHARGE Consortium (https://web.chargeconsortium.com/main/results). Summary statistics for intracranial aneurysms and aneurysmal subarachnoid hemorrhage can be found in the original literature (PMID: 33199917, PMID: 34594039) and the FinnGen database (https://www.finngen.fi/en/access_results). Additionally, summary statistics for the mediators are accessible from the GWAS catalog (https://www.ebi.ac.uk/gwas/).

https://www.finngen.fi/en/access_results

https://www.ebi.ac.uk/gwas/

https://doi.org/10.6084/m9.figshare.11303372.

## Acknowledgments

We extend our heartfelt gratitude to the ISGC Intracranial Aneurysm working group for providing the GWAS data on IAs and aSAH. Our sincere appreciation goes to the participants and investigators of several large-scale GWAS consortia, including the UK Biobank, Biobank Japan, FinnGen, CHARGE, GLGC, MAGIC, and the Blood Cell Consortium, for making their summary data available. We are also grateful to Prof. Surendran P., Prof. Ligthart S., Prof. Zhao JH., and their colleagues for providing GWAS datasets of mediators. Additionally, we thank Liwen Bianji (Edanz) (https://www.liwenbianji.cn) for their assistance in editing the English text of a draft of this manuscript.

## Author contributions

D.W., X.C., and Y.L. had full access to all the data in the study and take responsibility for the integrity of the data and the accuracy of the data analysis. D.W., X.C., S.G., M.L., F.M., and Y.L. conceived and designed the study. D.W. and X.C. undertook the statistical analyses. D.W. and S.G. made figures. D.W. wrote the first draft of the manuscript. All authors provided guidance on statistical analyses and visualization. Critical revisions for important intellectual content were made by all authors. The final version of the manuscript has been reviewed and approved by all authors.

## Ethical approval and informed consent statements

Ethical review and approval were waived for this study, as it utilized data from public databases. Ethical permits were obtained by the individual studies included in the analyses from their respective Local Ethical Committees. Informed consent was acquired from all subjects involved in the studies within the public database.

## Supplementary material

Tables S1-S30

Supplementary Methods

