## Supplementary Methods for "Omega-3 fatty acids and intracranial aneurysms: a Mendelian randomization study"

**Detailed information on sources of exposure and outcome**

1.UK Biobank

Between April 2007 and December 2010, UK Biobank enrolled 502,655 participants (5.5% response rate) aged 40–69 years from the general population. In June 2018, Nightingale Health released a Genome-Wide Association Study (GWAS) on plasma DHA concentrations measured by nuclear magnetic resonance spectroscopy in 114,999 individuals. We obtained summary-level data from this study as the exposure dataset and extracted instrumental variables for analysis.

2.Bakker et al.’s study

In 2020, Bakker et al. conducted a two-stage cross-ancestry genome-wide association study focusing on patients diagnosed with intracranial aneurysms in community or hospital settings. The first stage involved analyzing data from individuals of European ancestry, combining genotypes from 23 cohorts, comprising 7,495 cases and 71,934 controls, with analysis of 4,471,083 SNPs. In the second stage, they integrated data from two East Asian ancestry datasets, totaling 10,754 cases, 306,882 controls, and 3,527,309 SNPs. IA and aSAH data from both stages were utilized as outcome data. To avoid sample overlap, individuals from UK Biobank were removed from all datasets.

3.Sakaue et al.’s study

In 2021, Sakaue et al. conducted 220 deep-phenotype genome-wide association studies (GWAS) encompassing diseases, biomarkers, and medication usage in the Biobank Japan cohort (n = 179,000), integrating past medical history and text-mining of electronic medical records. We obtained a dataset from their study, comprising 3,132 patients of East Asian ancestry with documented intracranial aneurysm diagnoses in their medical history or electronic medical records, alongside 155,154 controls.

4.Finngen database

The FinnGen study, a large-scale genomics initiative, has scrutinized over 500,000 Finnish biobank samples, correlating genetic variation with health data to unravel disease mechanisms and predispositions. This collaborative endeavor involves research organizations, biobanks in Finland, and international industry partners. The FinnGen database provides comprehensive data on both unruptured and ruptured aneurysms. The unruptured aneurysm dataset comprises 2,784 patients and 374,631 controls, while the ruptured aneurysm dataset includes 5,754 patients with aneurysmal subarachnoid hemorrhage who underwent surgery, alongside 374,631 controls. We integrated these datasets as intracranial aneurysm outcome data using PLINK (version 1.90b7.2).

**Detailed introduction to Mendelian randomization**

Observational studies often suffer from various biases, such as reverse causality and confounding factors, which limit their ability to infer causal relationships. The instrumental variable (IV) method, commonly used in econometrics, is a technique to address endogeneity, which corresponds to confounding factors, reverse causality, and regression dilution bias in epidemiology. Mendelian Randomization (MR) uses genetic variants as IVs to infer causal relationships. The core principle of MR is to use instrumental variables that are not related to confounding factors in place of traditional exposures, thereby avoiding the influence of confounders.^1,2^ Moreover, using genetic variation avoids the issue of reverse causality, as genetic exposure is determined before the outcome occurs, and the outcome does not alter the genetic variant.

However, the selected genetic variants or instrumental variables must satisfy certain assumptions: the genotype is robustly associated with the modifiable (non-genetic) exposure of interest (Assumption 1); the genotype is not associated with confounding factors that bias conventional epidemiological associations between modifiable risk factors and outcomes (Assumption 2); and the genotype is related to the outcome only via its association with the modifiable exposure (Assumption 3). These assumptions ensure that statistical analysis can test whether the exposure affects the outcome. To obtain an interval estimate of the causal effect, it is also necessary to assume that the associations between the instrumental variable, exposure, and outcome are linear and statistically without interaction effects.^2^

We can use the Wald ratio estimator to estimate the regression coefficient for the effect of exposure (X) on outcome (Y) when there is only a single IV.^3^

$$\hat{\beta}_{\mathrm{IV}}={\hat{\beta}_{\mathrm{ZY}}}/{\hat{\beta}_{\mathrm{ZX}}}$$

where $\hat{\beta}_{\mathrm{ZY}}$ is the coefficient for the regression of the outcome (Y) on the IV (Z), and $\hat{\beta}_{\mathrm{ZX}}$ is the coefficient for the regression of exposure (X) on the IV (Z). The IV estimator $\hat{\beta}_{\mathrm{IV}}$ provides an estimate of the causal effect of exposure on the outcome.

The standard error of the Wald ratio is calculated as follows:

$$se\left( \hat{\beta}_{\mathrm{IV}} \right)=\left| {se(\hat{\beta}_{\mathrm{ZY}})}/{\hat{\beta}_{\mathrm{ZX}}} \right|$$

where $se\left( \hat{\beta}_{\mathrm{ZY}} \right)$ is the standard error for the regression of outcome (Y) on the IV (Z).

Numerous genome-wide association studies have identified tens or hundreds of independent genetic variants reaching the established genome-wide significance level, making it possible to infer causality using multiple SNPs. There are two advantages to using multiple SNPs for causal inference: first, it provides more precise estimates, improving statistical power; second, it may address the issue of horizontal pleiotropy.^2,4^ Assuming that the exposure affects the outcome and that SNPs directly influence only the exposure, we can expect that each SNP's effect on the outcome is proportional to its effect on the exposure. This proportional factor (causal effect) is consistent across SNPs, ensuring that their individual causal ratio estimates are homogeneous. The more SNPs that meet this expectation, the less likely it is that the SNP-outcome association is due solely to horizontal pleiotropy or different causal variants.^4,5^ We can use the inverse-variance weighted (IVW) method to combine the results of multiple genetic variants, which involves inverse variance weighting of the Wald ratios obtained from each genetic variant.^5^

Because the IVW estimate is essentially a weighted average of the Wald ratios obtained from each SNP, if any SNP exhibits horizontal pleiotropy (i.e., affects the outcome through pathways other than the exposure), the causal effect estimate may be biased.^4^ MR-Egger regression addresses this issue by allowing for a non-zero intercept, which represents an estimate of the directional pleiotropic effect.^6^ However, the statistical efficiency of MR-Egger is significantly lower than that of IVW.^4^ In addition to MR-Egger, more robust estimates can be obtained using methods such as the weighted median and weighted mode, which are designed under the assumption that some instrumental variables may be invalid.^7,8^ These methods provide a more reliable estimate of the causal effect by minimizing the impact of invalid instruments.

Two-sample MR is an improvement over the traditional single-sample MR approach. In two-sample MR, the coefficients for the regression of the outcome (Y) on the IV (Z) and the regression of exposure (X) on the IV (Z) come from different samples.^9^ This method increases the feasibility and cost-efficiency of MR studies while losing little power.^9^
